# Prevalence and Predictors of Depression among Training Physicians in China: A Comparison to the United States

**DOI:** 10.1101/2020.04.12.20049882

**Authors:** Lihong Chen, Zhuo Zhao, Zhen Wang, Ying Zhou, Xin Zhou, Hui Pan, Fengtao Shen, Suhua Zeng, Xinhua Shao, Srijan Sen, Weidong Li, Margit Burmeister

## Abstract

Resident physician training is associated with a substantial increase in depression in the United States, with rates increasing from about 4% before internship to 35% at least once during the first year of residency^1^.

Here, we sought to assess whether the rate of depression among residents in China are similar to their US counterparts and identify the common and differential predictors of depression in the two training systems. We assessed 1006 residents across three cohorts (2016-2019) at 16 affiliated hospitals of Shanghai Jiao Tong University and Peking Union Medical College. In parallel, we assessed three cohorts of 7028 residents at 100+ US institutions.

At the Chinese institutions, similarly, the proportion of participants who met depression criteria increased from 9% prior to residency to 35% at least once during the first year of residency (P<0.0001), an increase similar in magnitude to residents during internship in US institutions. Among factors assessed before residency, prior history of depression and depressive symptom score at baseline were common factors associated with depression during residency in both China and the US. In contrast, neuroticism and early family environment were strongly associated with depression risk in the US but not in China. Young age was a predictor of depression in China but not in the US sample. Among residency training factors, long duty hours and reduced sleep duration emerged as predictors of depression in both China and the US.

To gain insight into whether differences in personal predictors between the residents in China compared to the US residents were driven more by differences between cohorts, or by training system differences, we compared US residents of East Asian descent to other US and Chinese residents. We found that for most predictors (age, Neuroticism, early family environment), US residents of East Asian descent were more similar to other US residents than to the residents training in China.

Overall, the magnitude of depression increase and work-related drivers of depression were similar between China and the US, suggesting a need for system reforms, and that the types of effective reforms may be similar across the two systems.

## Introduction

The first year of physician residency training, internship, is one of the most stressful periods during a medical career, characterized by high work load, new responsibilities, and inconsistent and insufficient sleep^1^. A large meta-analysis including 54 studies across 16 countries found that depression increases 5-fold during internship, with 25-30% of interns fulfilling criteria for depression at any given time^1^. However, little is known about depression among residents in China, as no Chinese studies met criteria for inclusion.

In 2014, China introduced a standardized residency program, providing an opportunity to systematically assess depression among residents in China. The Intern Health Study is the largest study of depression among residents in the United States ^2,3^. Here, we longitudinally assess first-year resident physicians in China and the US utilizing the same Intern Health Study protocol in both settings to establish the prevalence and drivers of depression among residents in China, and compare to the prevalence and drivers in the US.

## Methods

### Participants

We assessed 3 cohorts of residents in China, enrolled in 2016-2019, in 16 hospitals in Shanghai and Beijing. About 2 weeks before start of residency, when the classes for each hospital are verified, an email invitation was sent to eligible first-year residents at participating hospitals. We invited 3666 first year residents, with 1664 (45%) agreeing to take part and completing the online baseline survey. As this first year of residency, unlike a US internship, is already specialized, we refer to this as the first year of residency throughout.

In parallel, we assessed 3 cohort of residents in the US, enrolled between 2016-2019, in 100+ institutions across the country. 6-8 weeks before the start of internship, an invitation email was sent to eligible first-year residents at participating US institutions. We invited 14,723 first year residents, with 8266 (56%) agreeing to take part and completing the online baseline survey online.

This study design and informed consent procedure was approved by the Ethics committees of Peking Union Medical College, Shanghai Jiao Tong University and the University of Michigan.

### Data collection

All surveys were conducted through a secure Web site or via a mobile app designed to maintain confidentiality, with subjects identified only by numbers. Depressive symptoms were measured at baseline and months 3, 6, 9, and 12 of their first year of residency, using the 9-item Patient Health Questionnaire (PHQ-9). The PHQ-9 is a self-report component of the Primary Care Evaluation of Mental Disorders inventory, designed to screen for depressive symptoms. For each of the 9 depressive symptoms, residents indicated whether, during the previous 2 weeks, the symptom had bothered them “not at all,” “several days,” “more than half the days,” or “nearly every day.” Each item yields a score of 0 to 3, so that the PHQ-9 total score ranges from 0 to 27.

#### Baseline Assessment

Subjects completed a baseline survey prior to commencing residency. The survey assessed general demographic factors (age, sex, ethnicity and marital status), medical education factors (medical institution and specialty), personal factors (baseline PHQ-9 depressive symptoms and self-reported history of depression) and the following psychological measures: (1) neuroticism (NEO-Five Factor Inventory), (2) early family environment (Risky Families Questionnaire). For first year residents in China, validated Chinese versions of inventories and translations non-inventory items were administered.

#### Within-Residency Assessments

Participants were contacted via e-mail at months 3, 6, 9, and 12 of their first year of residency and asked to complete the PHQ-9. They were also queried regarding their rotation setting, perceived medical errors, work hours, and sleep during the past week and the occurrence of a series of non-residency life stressors (serious illness; death or serious illness in a close family member or friend, financial problems; end of a serious relationship; or becoming a victim of crime or domestic violence) during the past 3 months. Chinese residents were also asked three questions about workplace violence, both at baseline and at each of the quarterly assessments: Have you experienced, observed or do you fear being physically assaulted, verbally abused, or threatened by a patient or their relatives?

### Statistical analysis

All analysis was performed using SAS version 9.4. P values less than 0.05 were considered statistically significant.

#### Prevalence of Depressive Symptoms and Depression Diagnostic Criteria

PHQ depression was defined by the criteria developed by Kroenke et al^4^, i.e. PHQ-9 score ≥ 10, which has a sensitivity of 93% and a specificity of 88% for the diagnosis of major depressive disorder.

To investigate whether there was a significant change in depressive symptoms or in depression prevalence during the first year of residency, we compared baseline PHQ-9 depressive symptoms and depressive symptoms at the 3-, 6-, 9-, and 12-month assessments through a series of paired t-tests. We also compared the percentage of subjects meeting diagnostic criteria for depression between baseline and during residency through a series of McNemar’s tests.

#### Predictors of Depression

To identify baseline variables that predict change in depressive symptoms during residency, we used Pearson correlations for continuous measures and Chi-square analyses for nominal measures. Significant variables were subsequently entered into a stepwise linear regression model to identify significant predictors while accounting for collinearity among variables. The association between variables measured through quarterly assessments during the first year of residency (within-residency variables) and change in depressive symptoms were assessed through a series of generalized estimating equation (GEE) analyses to account for correlated repeated measures within subjects. Variables (work hours, occurrence of medical errors, hours of sleep, non-residency stressful life events and, in China only, fear of workplace violence stressful events) were incorporated as predictor variables, and associated baseline factors were used as covariates.

The East Asian ancestry subset of the US cohort was composed of subjects who both self-reported East Asian ethnicity and had a genomic profile that clustered as East Asian^5^. Two sample t-tests were used to compare the PHQ depression scores between the samples from China, the US, the US-East Asian and the remainder of the US subsample (without US-East Asian). Chi-square analyses were used to compare the PHQ depression rates between the samples from China, the US, the US East Asian and the remainder of the US subsample. Baseline predictors and GEE modeling was also performed separately for these groups.

## Results

In China, 1057 (64%) subjects completed at least one follow-up survey and 1006 remained for analysis after deleting those missing gender and age. In the US, 7120 (86%) subjects completed at least one follow-up survey and 7028 remained for analysis.

### Prevalence of Depressive Symptoms

Among first year residents in China, the mean PHQ-9 depressive symptom score increased significantly from baseline (4.0±4.0) to 3 months (6.2±4.9; P<.0001), 6 months (6.5±5.2; P<.0001), 9 months (6.6±5.1; P<.0001), and 12 months (6.8±5.6; P<.0001) of residency. The increase is similar to that reported previously in the US^2^ and in the US sample reported here (Table 2). The mean PHQ9 score in China was significantly higher than the US score at baseline, and at 3, 6, 9 and 12 months of residency (Table 2).

**Table 1.**
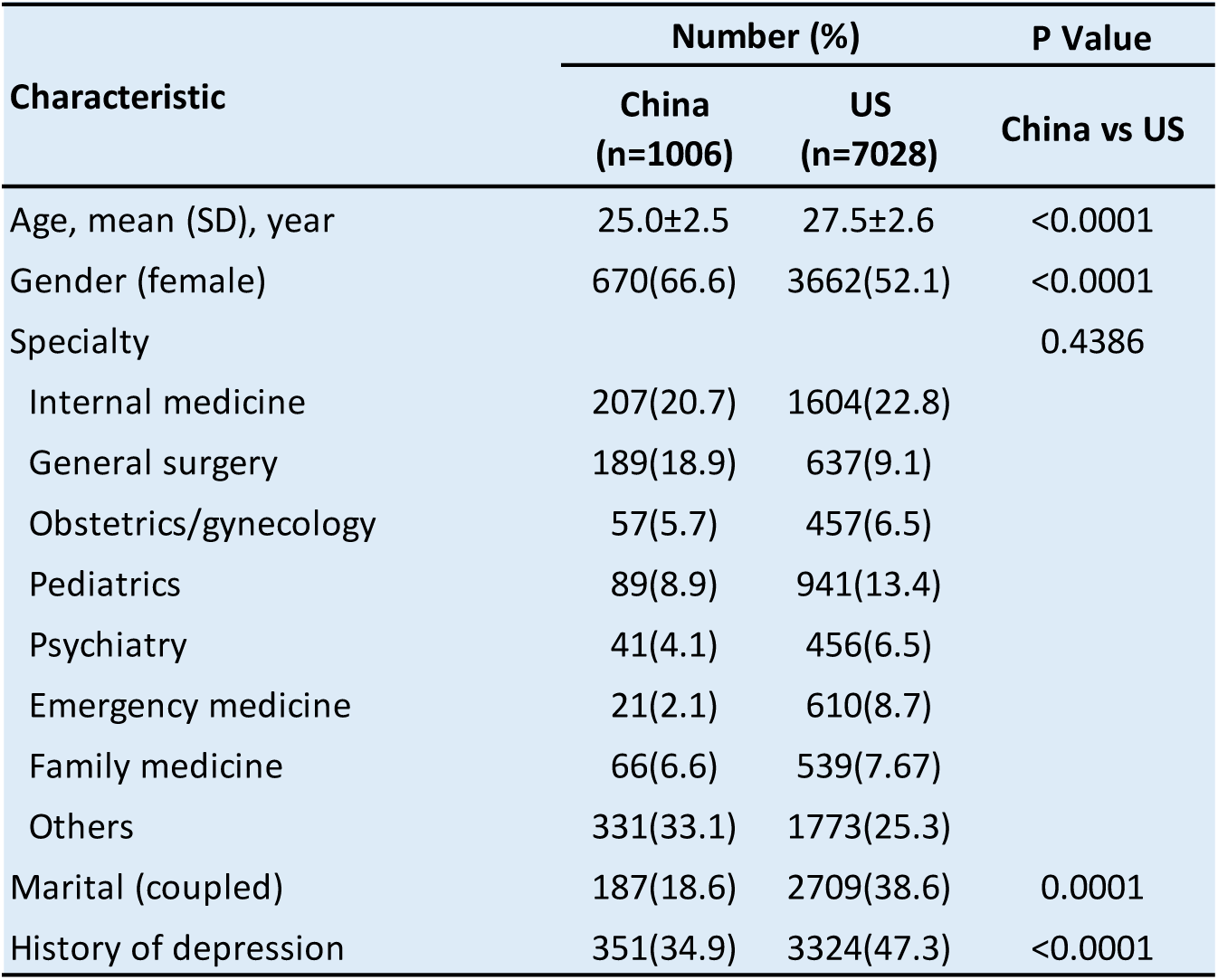
**Sample Demographic Characteristics at Baseline**

| Table 1 Sample Demographic Characteristics at Baseline |  |  |  |
| --- | --- | --- | --- |
| Characteristic | Number (%) |  | P Value |
|  | China<br>(n=1006) | US<br>(n=7028) | China vs US |
| Age, mean (SD), year | 25.0±2.5 | 27.5±2.6 | <0.0001 |
| Gender (female) | 670(66.6) | 3662(52.1) | <0.0001 |
| Specialty |  |  | 0.4386 |
| Internal medicine | 207(20.7) | 1604(22.8) |  |
| General surgery | 189(18.9) | 637(9.1) |  |
| Obstetrics/gynecology | 57(5.7) | 457(6.5) |  |
| Pediatrics | 89(8.9) | 941(13.4) |  |
| Psychiatry | 41(4.1) | 456(6.5) |  |
| Emergency medicine | 21(2.1) | 610(8.7) |  |
| Family medicine | 66(6.6) | 539(7.67) |  |
| Others | 331(33.1) | 1773(25.3) |  |
| Marital (coupled) | 187(18.6) | 2709(38.6) | 0.0001 |
| History of depression | 351(34.9) | 3324(47.3) | <0.0001 |

**Table 2.** PHQ-9 Depression Symptoms Score. Comparison of PHQ9 before and during first year of residency in China and US. The absolute PHQ9 scores at baseline and during 3 month intervals in residents in China and the US. The baseline and all scores are higher in China, but the difference (increase) during residency is less in China than in the US.

| Table 2 PHQ-9 Depression Symptoms Score |  |  |  |
| --- | --- | --- | --- |
|  | Mean (SD) |  | P Value |
|  | China | US | China vs US |
| Baseline | 4.0 ( 4.0 ) | 2.6 ( 3.0 ) | <0.0001 |
| 3 Months | 6.2 ( 4.9 ) | 5.7 ( 4.3 ) | 0.01 |
| 6 Months | 6.5 ( 5.2 ) | 6.0 ( 4.6 ) | 0.009 |
| 9 Months | 6.6 ( 5.1 ) | 5.9 ( 4.6 ) | 0.0027 |
| 12 Months | 6.8 ( 5.6 ) | 5.8 ( 4.7 ) | 0.0002 |
| Residency Average | 6.7 ( 4.6 ) | 6.0 ( 4.0 ) | <0.0001 |
| Change Score |  |  |  |
| 3 Months $\Delta$ | 2.3 ( 4.4 ) | 3.1 ( 4.0 ) | <0.0001 |
| 6 Months $\Delta$ | 2.6 ( 5.0 ) | 3.4 ( 4.2 ) | 0.0001 |
| 9 Months $\Delta$ | 2.9 ( 5.0 ) | 3.4 ( 4.2 ) | 0.0183 |
| 12 Months $\Delta$ | 3.0 ( 5.4 ) | 3.3 ( 4.4 ) | 0.1868 |
| Average $\Delta$ | 2.6 ( 4.4 ) | 3.4 ( 3.7 ) | <0.0001 |

Among residents in China, the percentage of subjects meeting diagnostic criteria for depression increased from 9.1% at baseline to 21.1%, 25.7%, 23.4%, and 28.0% at the 3-, 6-, 9-, and 12-month points of residency, respectively (Figure 1). 35.1% (353 of 1006) of subjects in China met criteria for major depression at least once during the first year of residency. In the US sample, the percentage of subjects meeting PHQ depression criteria increased from 3.9 % at baseline to 18.1%, 20.6%, 20.2%, and 20.1% at the 3-, 6-, 9-, and 12-month points of internship, respectively (Figure 1). 34.9% (2454 of 7028) of US subjects met criteria for major depression at least once during internship. Hence, the number of subjects meeting depression diagnostic criteria during the first year of residency is very similar between China and the US (35.1 vs 34.9%, p=0.91).

**Figure 1:**
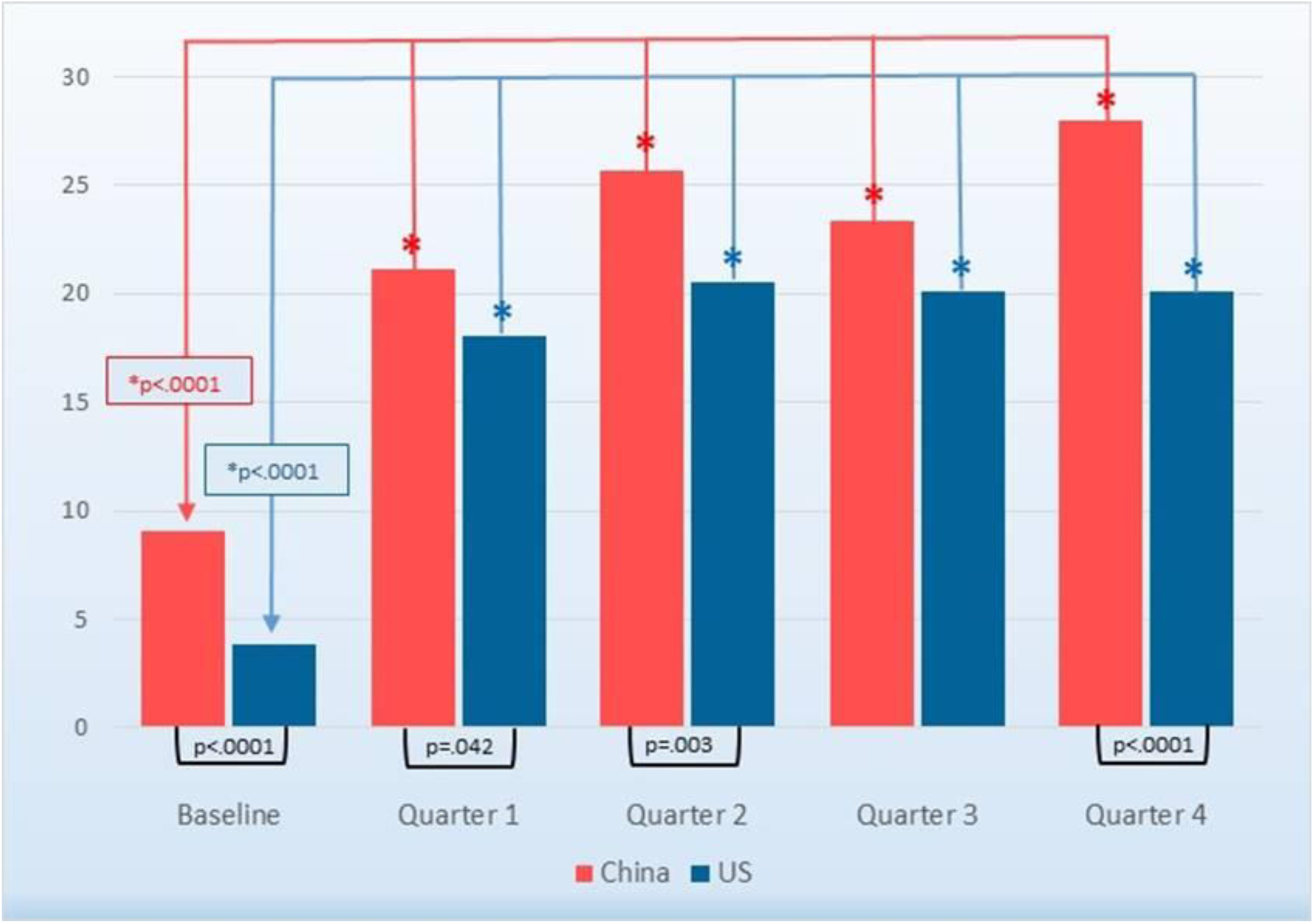
Depression Rates of Residents in China and the United States. The PHQ depression rate of residents is significantly higher in China (red) compared to the full US (blue) sample, at baseline and at the 1st, 3rd and 4th quarterly assessments (P value below histograms). Each 3-month assessment is significantly higher than the corresponding baseline PHQ-9 score (*p<0.0001, above the histograms).

### Baseline Predictors of Depression

Among 9 baseline variables tested among both Chinese and US residents, 3 were significant predictors of PHQ9 score change within residency in both China and the US: personal history of depression, stressful life events and a lower baseline depression symptom level (Table 3). Young age was predictive only in China, while female gender, neuroticism score, early family environment and not being coupled were predictive only in the US (Table 3). Workplace violence was only assessed in China. Fear of workplace violence was a significant predictor.

**Table 3.** Baseline Factors Correlation with Changing Depressive Symptoms. Baseline predictors of change in PHQ9 score during residency (average all 4 follow-ups) in China and the US. Significant correlations are bolded.

| **Table 3 Baseline Factors Correlation with Changing Depressive Symptoms** |  |  |  |  |
| --- | --- | --- | --- | --- |
| **Baseline Factors** | **China** |  | **US** |  |
|  | **r** | **P Value** | **r** | **P Value** |
| Age | **-0.1711** | **<0.0001** | -0.0012 | 0.9183 |
| Neuroticism | -0.0358 | 0.2577 | **0.1414** | **<0.0001** |
| Early family environment | -0.0089 | 0.7784 | **0.0919** | **<0.0001** |
| Baseline depressive symptoms | **-0.3899** | **<0.0001** | **-0.2757** | **<0.0001** |
| Gender (female) | 0.0408 | 0.2941 | **0.0758** | **<0.0001** |
| Marital (coupled) | 0.0682 | 0.0799 | **0.0486** | **0.0002** |
| Personal history of depression | **0.2278** | **<0.0001** | **0.2618** | **<0.0001** |
| Stressful life events | **0.1051** | **0.0069** | **0.0732** | **<0.0001** |
| Specialty | 0.0794 | 0.7605 | 0.0227 | 0.8757 |
| Fear of workplace violence | **0.2383** | **<0.0001** | NA | NA |

### Within-Residency Predictors of Depression

Using GEE analysis to control for repeated measures, we identified residency factors that were associated with increased depressive symptoms. Three within-residency factors were significantly associated with an increase in depressive symptoms in both countries: work hours, non-internship stressful life events and average sleep hours (Table 4). Reported medical errors were significantly associated only in the US. Experience, witnessing and fear of violence by patients or patients’ relatives against physicians was only measured in China. 8.2% of residents in China reported experiencing violence, while 24.1% reported witnessing and 17.4% feared violence. Fear of violence during residency was the best predictor and was retained in the GEE model.

**Table 4.** Predictors of Increased Depressive Symptoms during Residency. GEE model of significant baseline and within residency predictors in China and the US. Empty space – these factors were not originally significant or not measured, All significant factors in the final model are bolded.

| **Table 4 Predictors of Increased Depressive Symptoms during Residency** |  |  |  |  |
| --- | --- | --- | --- | --- |
| **Predictors** | **China** |  | **US** |  |
|  | **β** | **P Value** | **β** | **P Value** |
| **Baseline Factor** |  |  |  |  |
| Age | -0.2761 | <0.0001 |  |  |
| Gender (female) |  |  | 0.0802 | 0.2924 |
| Marital (coupled) |  |  | -0.2446 | 0.0009 |
| Neuroticism |  |  | 0.1213 | <0.0001 |
| Early family environment |  |  | 0.0327 | <0.0001 |
| Baseline depressive symptoms | 0.4914 | <0.0001 | 0.4122 | <0.0001 |
| Personal history of depression | 1.7059 | <0.0001 | 0.8989 | <0.0001 |
| **Residency Factor** |  |  |  |  |
| Work hours | 0.0331 | <0.0001 | 0.0428 | <0.0001 |
| Sleep hours | -0.3432 | <0.0001 | -0.2099 | <0.0001 |
| Medical errors | 0.1656 | 0.1324 | 0.8701 | <0.0001 |
| Stressful life events | 0.7037 | 0.0002 | 0.3423 | <0.0001 |
| Fear of workplace violence | 0.2043 | <0.0001 |  |  |

### Training factors rather than ethnicity explain most differences between the samples from China and the US

Since several baseline and within residency factors differed between the residents from China and those from the US, we wondered whether these differences were due to ethnicity, or due to differences in training structure. We took advantage of the fact that 13.7% of US trainees, N=963, were of East Asian ancestry, comparable in number to the residents in China (N=1006), and compared both the baseline and the within residency predictors to the US residents of other ancestry (Table 5).

**Table 5.** Predictors of Increased Depressive Symptoms during Residency. Comparison of significant baseline and residency factors between residents in China and US-residents of East Asian and those of non-East Asian descent.

|  | China (N=1006) |  | US-EastAsian (N=963) |  | US-nonEastAsian (N=6065) |  |
| --- | --- | --- | --- | --- | --- | --- |
| | $\beta$ | P Value | $\beta$ | P Value | $\beta$ | P Value |
| **Baseline Factor** |  |  |  |  |  |  |
| Age | **-0.2761** | **<0.0001** |  |  |  |  |
| Gender (female) |  |  | -0.1746 | 0.3740 | 0.1278 | 0.1215 |
| Marital (coupled) |  |  | **-0.4024** | **0.0423** | **-0.2478** | **0.0017** |
| Neuroticism |  |  | **0.1175** | **<0.0001** | **0.1233** | **<0.0001** |
| Early family environment |  |  | **0.0222** | **0.0302** | **0.0359** | **<0.0001** |
| Baseline depressive symptoms | **0.4914** | **<0.0001** | **0.3489** | **<0.0001** | **0.4189** | **<0.0001** |
| Personal history of depression | **1.7059** | **<0.0001** | **1.1599** | **<0.0001** | **0.8355** | **<0.0001** |
| **Residency Factor** |  |  |  |  |  |  |
| Work hours | **0.0331** | **<0.0001** | **0.0459** | **<0.0001** | **0.0423** | **<0.0001** |
| Sleep hours | **-0.3432** | **<0.0001** | **-0.2423** | **<0.0001** | **-0.2051** | **<0.0001** |
| Medical errors | 0.1656 | 0.1324 | **0.9748** | **<0.0001** | **0.8540** | **<0.0001** |
| Stressful life events | **0.7037** | **0.0002** | **0.6219** | **0.0012** | **0.2965** | **<0.0001** |

For most baseline and residency factors that differed between residents from US and China, the East Asian US residents were more similar to the other US residents (Table 5), suggesting that the differences observed were mainly due to the structure of the residency in China compared to the US. Two exception were female gender and marital/coupled status. Female gender was associated with higher PHQ depression in the US sample on its own, but not in the final model accounting for all factors, and not in the sample from China. Being married or coupled was not a predictor in China or in the East Asian US subsample, but that may be due to the small number of coupled residents due to the younger age in China. Hence, overall, there is remarkable consistency.

## Discussion

This study is the first to assess depression during the first year of residency training in China. Despite structural differences, we find striking parallels in prevalence and system predictors of depression between China and the US, with about 35% of participants fulfilling criteria for depression at least once during the first year of residency in both systems. In addition, while most baseline and within residency predictors were common between countries, interesting differences were also emerged.

Notably, this is the first large scale study to establish the level of depression in China under the new standardized training system. The finding of high prevalence indicates that depression among training physicians is a major problem in China, as it is in the rest of the studied world. The finding that the most system level predictors of depression are common between China and the US suggest that successful interventions developed in one system may succeed in the other system.

In contrast to the similar levels of depression during the first year of residency, the prevalence of PHQ depression measured at baseline, before the start of internship was substantially higher among training physicians in China. This baseline difference may be attributable to the difference in length of the low stress period that typically precede residency in the two systems: Specifically, in the US, there is typically a 2-3 month break between the end of intense medical school rotations and the start of internship. By contrast, Chinese residents have on average only 2 weeks between the end of medical school and start of residency.

We identified several individual baseline factors associated with the development of depressive symptoms during residency. Consistent with previous studies^1^, a history of major depression and of other stressful life events were common predictors in both samples. Higher baseline PHQ9 scores predicted higher overall PHQ9 scores during residency, but predicted smaller increases during residency compared to baseline in both samples.

Residency in China starts most commonly after 5 years of a Bachelor in Medicine, while in the US, 4 years of undergraduate studies are followed by 4 years of medical school. Hence, the Chinese sample was on average 2.5 years younger than the US sample, which also resulted in fewer Chinese residents compared to US interns being married or living together with a partner. Younger age was a significantly predictor of depression in China but not in the older US sample. A subset (about 15%) of the Chinese sample had finished an MS or PhD prior to residency, and hence were older. This small group reported lower depression rates compared to those starting right after the Bachelor (data not shown). In the final prediction model, once age was considered, the type of resident (BS, MS or PhD prior to residency) was not significant, but not vice versa, indicating that while age and type of resident are confounded, age is the dominant factor. Consistent with the literature^1^, female gender and not being in a relationship were predictors of depression in the US sample. Gender was not a significant predictor in China or among the East Asian US residents.

The personality trait, neuroticism is the strongest and among the best replicated predictors of depression during residency in the US^2,5^. In addition to the overall US sample (Table 1), neuroticism was also a strong predictor in the subsample of East-Asian ethnicity in the US (Table 5). In striking contrast, neuroticism has no predictive power for depression for residents in China. In fact, the non-significant trend of the neuroticism-depression association was in the opposite direction. This finding supports previous work suggesting that anxiety-related personality traits serve as vulnerability factors for disease in individualistic cultures, such as the US, but serve as protective factors in collectivist cultures such as China^6,7^.

Consistent with previous studies^1,2^, reduction of sleep,^8^ longer work hours^2,9^ and stressful events outside of residency^2^ were robust predictors of the observed increase in depression during residency in both countries. Notably, residency specialty was not associated with the development of depression in either country.

Another striking difference between the cohorts was the frequency of self-reported medical errors. While almost 20% of US residents reported errors each quarter, less than 2% of Chinese residents reported errors. This difference may be due to a number of potential factors including differences in the level of supervision or training between the two systems and differences the definition of errors or willingness to admit errors. Violence by patient or patients’ relatives against providers has recently been much discussed^10^, and fear of such violence was a significant predictor in the final model of depression during residency in China.

Our study has a number of limitations. First, since only about half of residents completed our questionnaires, it is conceivable that those who are suffering of depression symptoms during residency are more motivated to complete assessments. Second, we were only able to include residents from large hospitals in Shanghai and Beijing, who likely are not representative of training physicians across China. Finally, we focused on first-year residents in this study. Prevalence and predictors of depression may be different in more advanced training physicians.

In summary, similar to the US, the first year of residency in China is a stressful time leading to a marked increase of depression. While individual risk and protective factors differ between the countries, high work load and lack of sleep are major modifiable factors increasing risk for depression in both countries during the first year of residency.

## Data Availability

Data will be available upon request as long as legally and ethically appropriate.

## Acknowledgement

We thank all the residents in both countries for participation, and all hospital residency directors for participating in our study. This study was funded in China by Shanghai Jiao Tong University-University of Michigan Collaborative Research Grant, Program of Shanghai Subject Chief Scientist 17XD1401700, Shanghai Education Commission Research and Innovation Program, “Eastern Scholar” and “111” Project.

